# GALEAS™ Bladder Demonstrates High Sensitivity and Specificity for Detecting Bladder Cancer: Real-World Multicentre Data from UK NHS Haematuria Clinics

**DOI:** 10.64898/2026.02.06.26345752

**Authors:** Lee Silcock, Robert K Hastings, Samuel Clokie, Rebecca Sadler, Michael Parks, Matthew Smith, Victoria Hewitt, Jayne Louise Douglas-Moore, Helena Wignall, Hugo Escabelado, John Piedad, Sagar Kanabar, Chris Blick, Amar Mohee, Niall Pumfrey, Nadine Coull, Andy Goffe, Richard Tippett, Alexander Laird, Chandrarajan Premal Shah, Murray MacKay, Catherine Owen, Uwais Mufti, Douglas G Ward, Richard T Bryan

## Abstract

**Background:** Cystoscopy is a core component of haematuria investigations but is invasive and resource-intensive. GALEAS™ Bladder is a DNA-based diagnostic urine test that measures alterations in 23 bladder cancer-associated genes.

**Objective:** To assess the diagnostic performance and clinical utility of GALEAS™ Bladder as a molecular triage tool in real-world haematuria investigation pathways.

**Methods:** Patients referred for urgent investigation of haematuria were prospectively enrolled across seven UK NHS Urology Departments between October 2024 and June 2025. Urine samples were collected prior to cystoscopy and analysed using the GALEAS™ Bladder assay (Nonacus Clinical Services, UK). Assay results were compared with cystoscopy findings.

**Key Findings and Limitations:** Cystoscopic findings and GALEAS™ Bladder results were available for 964 participants, including 77 (8.0%) newly-diagnosed with pathology-confirmed BC. The assay demonstrated an overall sensitivity of 92.2% (95% CI: 84.0-96.4%), specificity of 92.0% (95% CI: 90.0-93.6%), and negative predictive value (NPV) of 99.3% (95% CI: 98.4-99.7%) for the diagnosis of BC. For the diagnosis of high-grade BCs, sensitivity was 97.2% (95% CI: 85.8-99.5%) with an NPV of 99.9% (95% CI:99.3-100.0%). Limitations include an absence of subsequent diagnoses for participants with positive GALEAS™ Bladder test results in the absence of cystoscopically-visible tumour.

**Conclusions and Clinical Implications:** GALEAS™ Bladder is a clinically implementable molecular urine test with very high sensitivity and specificity for the diagnosis of new cases of BC in patients undergoing urgent investigation of haematuria, especially for high-grade BCs. Clinical adoption could permit the molecular triage of haematuria patients to immediate or deferred cystoscopy.

## INTRODUCTION

Bladder cancer (BC) ranks among the ten most common malignancies globally, with over 18,000 new cases diagnosed annually in the UK and approximately 550,000 worldwide [1,2]. While haematuria is present in more than 90% of patients diagnosed with BC, only 8-19% of haematuria referrals are ultimately diagnosed with BC [3,4]. Flexible cystoscopy and urinary tract imaging, combined with histopathological analysis of resected tissue, remains the gold standard for BC detection [5] but is invasive, requires in-person attendance, and is subject to operator-dependent variability, with reported sensitivity and specificity for diagnostic cystoscopy of 81% and 73%, respectively [6]. Consequently, there is strong demand for accurate non-invasive diagnostic alternatives [7].

GALEAS™ Bladder is a Next Generation Sequencing (NGS)-based assay that employs a multi-marker panel and analytical module designed to detect a broad spectrum of BC-associated genetic mutations at low Variant Allele Frequency (VAF) in DNA extracted from urine samples [8,9]. Validation studies have demonstrated promising clinical sensitivity in both the haematuria (diagnosis) and surveillance (follow-up) settings, showing strong concordance with standard-of-care diagnostics [8]. We assessed the performance of GALEAS™ Bladder in UK NHS patients referred for urgent investigation following presentation with haematuria. Test results were compared against final diagnoses confirmed by cystoscopy and histological analysis.

## METHODS

### Design

We conducted an observational real-world evaluation study conducted across seven UK NHS Urology Departments. All participants underwent standard diagnostic procedures for haematuria investigation, including flexible cystoscopy for direct bladder visualisation, imaging to assess the upper urinary tract, and histological confirmation where indicated.

### Study Population

Eligibility criteria included adults aged ≥18 years referred for investigation following the detection of either visible haematuria (VH) or non-visible haematuria (NVH), with the ability to provide informed consent and a voided urine sample prior to cystoscopy. Exclusion criteria comprised a prior diagnosis of urothelial cancer, inability to provide an adequate urine sample, or unwillingness or inability to undergo standard haematuria investigations.

### Ethics

The study design was evaluated and approved at each institutions’ ethics committees and individual consent was obtained before samples were collected (where required by individual units) in accordance with the Declaration of Helsinki [10]. Demographic data, relevant medical history, and risk factors were collected by participating NHS centres. Outcomes from cystoscopy and histological analysis were recorded where both were available.

### Sample processing with GALEAS™ Bladder

All referrals, sample receipt, processing, and reporting were conducted at Nonacus Clinical Services (NCS), an ISO 15189 UKAS-accredited laboratory. Urine samples (≥25 ml) were collected primarily at home using the GALEAS™ Urine Collection Device (UCD); collection time was not recorded. The UCD contains 3.5ml preservative agent to inhibit bacterial growth and preserve DNA (final preservative:urine ratio 1:7-1:14). Samples were stored at room temperature for ≤28 days prior to processing. UCD content was centrifuged at 1500g for 10 minutes to isolate a pellet from which genomic DNA was extracted using the GALEAS™ Bead Xtract: Urine gDNA 96 kit [11]. DNA quantification was performed using Qubit (Thermo, USA). DNA libraries were prepared using the GALEAS™ Bladder NGS kit (Nonacus Ltd, UK), targeting a panel of >450 single nucleotide variants across 23 genes [8,9]. Sequencing was performed on an Illumina NextSeq 2000 (Illumina, USA) and FASTQ files processed using the proprietary GALEAS™ Bladder Software pipeline (Supplemental Methods).

### Statistical Analysis

This study aimed to recruit 1,200 patients, based on an estimated BC prevalence of 10.5% [3,4,12,13]. Accounting for an anticipated combined 10% participant dropout rate and patient non-compliance. This sample size was expected to yield approximately 113 BC positive and 967 BC-negative evaluable cases. See Supplemental Methods for additional calculations. Diagnostic performance metrics were calculated from 2×2 contingency tables. Likelihood ratios were computed as LR+ = sensitivity/(1-specificity) and LR-= (1-sensitivity)/specificity, with 95% confidence intervals via the Wilson score method. The diagnostic odds ratio (DOR) was calculated as (TP×TN)/(DP×FN), representing the odds of a positive test result in patients with cancer relative to those without (Supplementary Methods, Supplemental Table S3A and S3B).

## RESULTS

### Patient Demographic and Diagnosis Data

GALEAS Bladder test results with a final diagnosis were obtained for 964 participants enrolled between October 2024 and June 2025 from seven outpatient haematuria clinics in Urology Departments across seven UK NHS sites (Supplemental Table S1).

Patient demographics and histopathology confirmed diagnoses are summarised in Table 1; these characteristics are consistent with previously-reported haematuria cohorts [4,12–14]]. Bladder cancer was diagnosed in 77 patients (8.0%), with a median age of 77 years (IQR: 70-81) versus 66 years (IQR: 54-76) in patients with a non-BC diagnosis. VH was present in 575 patients (59.6%), of whom 68 (11.8%) were diagnosed with bladder cancer, compared to 7 of 294 (2.4%) patients with NVH.

**Table 1:** Patient Demographics in the real world multi centre NHS Study. Characteristics of participants stratified by bladder cancer diagnosis status. Continuous variables are reported as median (Q1, Q3); categorical variables as n (%). MIBC = muscle-invasive bladder cancer; NMIBC = non-muscle invasive bladder cancer. Haematuria type classified as visible (macroscopic) or non-visible (microscopic). T (Tumour) stage reported according to TNM classification; grade is reported according to WHO 2004-2016 classification.

| Characteristic | Overall N = 964<sup>1</sup> | Diagnosis |  |  |
| --- | --- | --- | --- | --- |
|  |  | MIBC N = 17<sup>1</sup> | NMIBC N = 60<sup>1</sup> | No Cancer N = 887<sup>1</sup> |
| Age | 67.0 (54.5, 76.0) | 74.0 (72.0, 79.0) | 77.5 (69.5, 81.0) | 66.0 (54.0, 76.0) |
| Sex |  |  |  |  |
| Male | 568 (58.9%) | 16 (94.1%) | 45 (75.0%) | 507 (57.2%) |
| Female | 391 (40.6%) | 1 (5.9%) | 14 (23.3%) | 376 (42.4%) |
| Not Provided | 5 (0.5%) | 0 (0.0%) | 1 (1.7%) | 4 (0.5%) |
| Haematuria Type |  |  |  |  |
| Visible | 575 (59.6%) | 16 (94.1%) | 52 (86.7%) | 507 (57.2%) |
| Non-visible | 294 (30.5%) | 1 (5.9%) | 6 (10.0%) | 287 (32.4%) |
| Not Provided | 95 (9.9%) | 0 (0.0%) | 2 (3.3%) | 93 (10.5%) |
| Tumour stage* |  |  |  |  |
| Ta | 40 (51.9%) | 0 (0.0%) | 40 (66.7%) | NA |
| T1 | 13 (16.9%) | 0 (0.0%) | 13 (21.7%) | NA |
| T2 | 8 (10.4%) | 8 (47.1%) | 0 (0.0%) | NA |
| T2a | 3 (3.9%) | 3 (17.6%) | 0 (0.0%) | NA |
| T2b | 3 (3.9%) | 3 (17.6%) | 0 (0.0%) | NA |
| T3 | 1 (1.3%) | 1 (5.9%) | 0 (0.0%) | NA |
| T4b | 1 (1.3%) | 1 (5.9%) | 0 (0.0%) | NA |
| Not Provided | 8 (10.4%) | 1 (5.9%) | 7 (11.7%) | NA |
| Tumour grade* |  |  |  |  |
| High Grade | 36 (46.8%) | 12 (70.6%) | 24 (40.0%) | NA |
| Low Grade | 19 (24.7%) | 0 (0.0%) | 19 (31.7%) | NA |
| Not Provided | 22 (28.6%) | 5 (29.4%) | 17 (28.3%) | NA |

<sup>1</sup>Continuous: median (Q1, Q3); categorical: n (%). \*Tumour stage and grade not applicable for No Cancer patients (shown as NA).

### Clinical Sensitivity and Specificity

Of the 77 diagnosed with BC by cystoscopy, there were 60 NMIBCs and 17 MIBCs. GALEAS™ Bladder correctly identified 71 BC cases (6 discordant negatives - all NMIBC) and correctly classified 816 of 887 non-BC samples as negative (71 discordant positives) (Table 2).

**Table 2.** Diagnostic performance of GALEAS™ Bladder stratified by diagnosis, tumour stage, tumour grade, and haematuria type. Performance metrics of GALEAS™ Bladder compared to cystoscopy and histopathology across 964 patients.

| Subgroup | N | TP<sup>1</sup> | FN | TN | DP | Sensitivity %<br>(95% CI)<sup>2</sup> | Specificity %<br>(95% CI)<sup>3</sup> | PPV %<br>(95% CI) | NPV % (95%<br>CI) |
| --- | --- | --- | --- | --- | --- | --- | --- | --- | --- |
| **Overall** |  |  |  |  |  |  |  |  |  |
| Overall | 964 | 71 | 6 | 816 | 71 | 92.2 (84.0–96.4) | 92.0 (90.0–93.6) | 50.0 (41.9–58.1) | 99.3 (98.4–99.7) |
| **By Diagnosis** |  |  |  |  |  |  |  |  |  |
| NMIBC | 60 | 54 | 6 | 0 | 0 | 90.0 (79.9–95.3) | 92.0 (90.0–93.6) | 43.2 (34.8–52.0) | 99.3 (98.4–99.7) |
| MIBC | 17 | 17 | 0 | 0 | 0 | 100.0 (81.6–100.0) | 92.0 (90.0–93.6) | 19.3 (12.4–28.8) | 100.0 (99.5–100.0) |
| No Cancer | 887 | 0 | 0 | 816 | 71 | — | 92.0 (90.0–93.6) | 0.0 (0.0–5.1) | 100.0 (99.5–100.0) |
| **By Stage** |  |  |  |  |  |  |  |  |  |
| Ta stage | 40 | 34 | 6 | 0 | 0 | 85.0 (70.9–92.9) | 92.0 (90.0–93.6) | 32.4 (24.2–41.8) | 99.3 (98.4–99.7) |
| T1 stage | 13 | 13 | 0 | 0 | 0 | 100.0 (77.2–100.0) | 92.0 (90.0–93.6) | 15.5 (9.3–24.7) | 100.0 (99.5–100.0) |
| T2+ stage<sup>4</sup> | 16 | 16 | 0 | 0 | 0 | 100.0 (80.6–100.0) | 92.0 (90.0–93.6) | 18.4 (11.6–27.8) | 100.0 (99.5–100.0) |
| **By Grade** |  |  |  |  |  |  |  |  |  |
| High Grade | 36 | 35 | 1 | 0 | 0 | 97.2 (85.8–99.5) | 92.0 (90.0–93.6) | 33.0 (24.8–42.4) | 99.9 (99.3–100.0) |
| Low Grade | 19 | 16 | 3 | 0 | 0 | 84.2 (62.4–94.5) | 92.0 (90.0–93.6) | 18.4 (11.6–27.8) | 99.6 (98.9–99.9) |
| **By Haematuria** |  |  |  |  |  |  |  |  |  |
| Visible Haematuria | 575 | 62 | 6 | 456 | 51 | 91.2 (82.1–95.9) | 89.9 (87.0–92.3) | 54.9 (45.7–63.7) | 98.7 (97.2–99.4) |
| Non-visible Haematuria | 294 | 7 | 0 | 271 | 16 | 100.0 (64.6–100.0) | 94.4 (91.1–96.5) | 30.4 (15.6–50.9) | 100.0 (98.6–100.0) |

<sup>1</sup>TP = true positive; FN = false negative; TN = true negative; DP = discordant positive; PPV = positive predictive value; NPV = negative predictive value.
<sup>2</sup>Performance metrics calculated with Wilson score 95% confidence intervals.
<sup>3</sup>For cancer-only subgroups (Diagnosis, Stage, Grade), Specificity uses overall population. PPV and NPV are calculated using subgroup TP/FN combined with No Cancer TN/DP.
<sup>4</sup>T2+ includes T2, T2a, T2b, T3, and T4b stages.

Tumour stage and grade for confirmed BC cases were assigned according to histopathology. The BC cohort comprised: 18 LG Ta, 13 HG Ta, 10 HG T1, 16 T2+, 9 Ta with grade not provided, 3 T1 with grade not provided, and 8 cases with stage not provided (Table 2).

The overall assay sensitivity, specificity, PPV and NPV was 92.2% (95%CI 84.0-96.4), 92.0% (95%CI 90.0-93.6), 50.0% (95%CI 41.9-58.1) and 99.3% (95%CI 98.4-99.7) respectively. Performance by subgroup is detailed in Table 2. For MIBC (n=17), sensitivity was 100% (95%CI 81.6-100), specificity 92.0% (95%CI 90.0-93.6), PPV 19.3 (95%CI 12.4–28.8) and NPV 100% (95%CI 99.5–100.0). For NMIBC (n=60), sensitivity was 90.0% (95%CI 79.9-95.3), specificity 92.0% (95%CI 90.0-93.6), PPV 43.2% (95% CI 34.8– 52.0), and NPV 99.3% (95%CI 98.4-99.7). For participants presenting with VH (n=575, 68 cancers), sensitivity was 91.2% (95%CI 82.1-95.9), specificity 89.9% (95%CI 87.0-92.3), PPV 54.9% (95%CI 45.7-63.7), and NPV 98.7% (95%CI 97.2-99.4). The NVH group (n=294, 7 cancers) achieved sensitivity of 100% (95%CI 64.6-100), specificity 94.4% (95%CI 91.1-96.5), PPV 30.4% (95%CI 15.6-50.9), and NPV 100% (95%CI 98.6-100).

GALEAS™ Bladder detected 35 of 36 high-grade bladder cancers (97.2% 95% CI: 85.8–99.5%) and 16 of 19 low-grade cancers (84.2%, 95% CI: 62.4–94.5%). Of the 6 discordant negatives, all were stage Ta (3 low-grade, 1 high-grade, and 2 unknown grade); 4 of the 6 measured <3cm in diameter. All MIBCs were detected (100%, 17/17) (Table 2).

If GALEAS™ Bladder were to be used as a triage test for the cystoscopic component of haematuria assessment, then the number of cystoscopies needed to diagnose one BC would reduce from 12.5 to 2.0, an 85.3% reduction. The positive likelihood ratio (LR+) was 11.5 for a GALEAS™ Bladder positive test and the negative likelihood ratio (LR-) was 0.085 for a negative test [15,16] The diagnostic odds ratio (DOR) was 136 (95% CI: 57.1–323.9). (Supplementary Methods, Supplemental Table S3).

### GALEAS™ Bladder molecular results

Among samples profiled, 142 harboured at least one BC-associated mutation, with a total of 285 mutations (mean 2.0 variants per positive sample; 54% (76/142) contained more than one alteration), Supplemental Table 4a. VAFs ranged from 0.2% to 85.2% (median 5.6%) (Supplemental Figures S2A, S2B).

Among the 71 true-positive samples, *TERT* promoter mutations were the most frequent, detected in 69.0% of samples. *FGFR3* and *PIK3CA* mutations were the next most common, present in 36.6% and 35.2% of samples, respectively. *ELF3* and *ERCC2* mutations were each observed in 14.1%. All detected mutations by grade and stage of disease are shown in Supplementary Table S4A and S4B.

Additional recurrent alterations occurred at lower frequencies in a broad set of canonical bladder cancer drivers (Supplemental Figure S2); both MIBC and NMIBC cases show broad VAF distributions with many high-fraction mutations.

Among the 71 discordant-positive samples (Table 2), *TERT* promoter mutations were the most frequently detected (56.3%), followed by *FGFR3* (11.3%) (Supplemental Figure S2). Within this discordant-positive group, we observed a bimodal distribution: 44/110 variants (40%) called were <1% VAF and 66/110 variants (60%) >1% VAF (Supplemental Figure S2).

## DISCUSSION

This real-world multicentre evaluation of GALEAS™ Bladder has demonstrated a diagnostic sensitivity of 92.2% (CI 84.0-96.4) and specificity of 92.0% (CI 90.0-93.6) for the detection of BC in typical UK haematuria patients. The assay showed consistent performance across all stages of bladder cancer, detecting 90.0% of NMIBCs and 100% of MIBCs. Furthermore, 97.2% of high-grade BCs were identified. Among the BC cases presenting with VH, GALEAS™ Bladder correctly identified 91.2%; all BC cases presenting with NVH were correctly identified (100% sensitivity).

The relevance of the study is illustrated by the cohort demographics: VH accounted for 88.3% of BC diagnoses, NVH accounted for 9.1%, and 2.6% had an unknown haematuria status. NMIBC represented 77.9% of diagnoses and MIBC 22.1%, with MIBC cases almost exclusively presenting with VH (94.1%). In contrast, NVH participants had a low BC incidence of 2.4%, including only a single case of MIBC.

These data build upon our previous studies [8,9], underscoring the reproducibility and clinical reliability of GALEAS™ Bladder as a diagnostic molecular urine test. The high NPV of 99.3% (98.7% for VH and 100% for NVH) means that the test is highly effective in ruling out BC in patients presenting with haematuria. The high positive (11.5) and low negative (0.085) Likelihood Ratios and a diagnostic odds ratio of 136 reinforce a role for GALEAS™ Bladder in triaging symptomatic patients for urgent or non-urgent cystoscopy as part of the suite of investigations required for the assessment of haematuria. Ultimately, this could lead to a >6-fold reduction in the number of urgent cystoscopies undertaken in this setting, thereby reducing waiting times.

The mutational spectrum in this RWE cohort mirrors the canonical genomic landscape of BC described by large-scale sequencing studies [9,19,20], with the frequent identification of *TERT* promoter mutations in 62.7% of positive cases (c.-124C>T accounting for the majority), *FGFR3* hotspot mutations in 38% (principally c.746C>G (p.S249C)), *PIK3CA* substitutions in 35% (p.E542K, p.E545K); mutations in *TP53* and *ERCC2* reflected the genomic instability characteristic of high-grade and muscle-invasive disease [19].

It is one of the limitations of the study that we did not systematically follow-up all patients to enquire about subsequent cancer/urothelial cancer diagnoses for those participants who had negative SOC investigations for the investigation of haematuria. Notwithstanding, anecdotal clinical feedback has indicated a number of subsequent urothelial cancer diagnoses in participants with positive GALEAS™ Bladder tests but negative SOC tests. As previously described [8], there is the potential for molecular detection below visual detection thresholds and the possibility of GALEAS™ Bladder detecting BC earlier than cystoscopy in some cases.

Other study limitations include the UK-based nature of the cohort; although a multicentre study across various devolved nations of the UK, it remains a study within the UK’s NHS. The absence of histological variants of BC within the cohort highlights the need to assess GALEAS™ Bladder in such specifically curated research cohorts. Nevertheless, study strengths include the prospective multicentre design, large sample size, real-world clinical setting, and ISO 15189 UKAS-accredited laboratory processing ensuring regulatory compliance.

GALEAS™ Bladder’s performance compares favourably to other NGS-based urine tests for BC detection. In comparable haematuria cohorts, UroSEEK demonstrated 83% sensitivity and 93% specificity [20], UroScout showed 96% sensitivity and 89% specificity [21], and UroAmp achieved 68-95% sensitivity (depending on stage) and 90% specificity [22]. Incorporating clinical risk prediction alongside molecular urine testing could further enhance triage for cystoscopy. For example, although not implemented in the UK, the IDENTIFY risk calculator [23] could be combined with GALEAS™ Bladder to identify very low composite risk patients.

In clinical practice, GALEAS™ Bladder is intended to inform pre-cystoscopy decision-making rather than replace standard diagnostic procedures. A negative result indicates a very low probability of clinically-significant disease (reflected by the high NPV and very low LR-) and supports deferral of immediate cystoscopy in NVH patients, while maintaining physician-directed surveillance. Conversely, a positive result supports prioritisation for prompt cystoscopy and histopathological confirmation. This risk-stratified approach enables more effective allocation of cystoscopy capacity by accelerating access for patients with a high probability of harbouring clinically-significant disease, while safely deferring procedures in low-risk patients; as demonstrated in the BladderPath trial [24], reducing time to correct treatment by as little as 6-weeks can improve cancer outcomes [25]. Therefore, by preserving near-complete detection of clinically-aggressive disease whilst potentially reducing urgent cystoscopy burden, GALEAS™ Bladder may provide direct clinical, health-system, and environmental benefit as a molecular triage tool within established haematuria diagnostic pathways.

## CONCLUSION

GALEAS™ Bladder is a clinically implementable molecular urine test with very high sensitivity and specificity for the diagnosis of new cases of BC in patients undergoing urgent investigation of haematuria. Clinical adoption could permit the triage of haematuria patients to immediate or deferred cystoscopy based upon urinary tumour DNA findings, thereby accelerating the diagnosis and subsequent treatment of the vast majority BCs.

## Data Availability

All data produced in the present study are available upon reasonable request to the authors

## ACKNOWLEDGEMENTS

The authors gratefully acknowledge the clinical team at Ward 28a, University Hospitals of Leicester NHS Trust, for their invaluable contributions to patient recruitment and sample collection. Abigail Coe, Emeka Udeh, Yaamini Premakular, Jin Tan, Alexander Lloyd and Giovanni Chiriaco from Kingston & Richmond NHS Foundation Trust. We thank Simon Davis, Saffron Kular, Myah Mahal and Elizabeth Coakley at Nonacus Ltd for project coordination and technical support, and the dedicated laboratory staff at Nonacus Clinical Services for sample processing and analysis. Charlotte Rich-Griffin, Jinli Luo and the Nonacus Bioinformatics team for data processing and analysis. We are deeply grateful to all patients who participated in this study.

---

**Figure 1.**
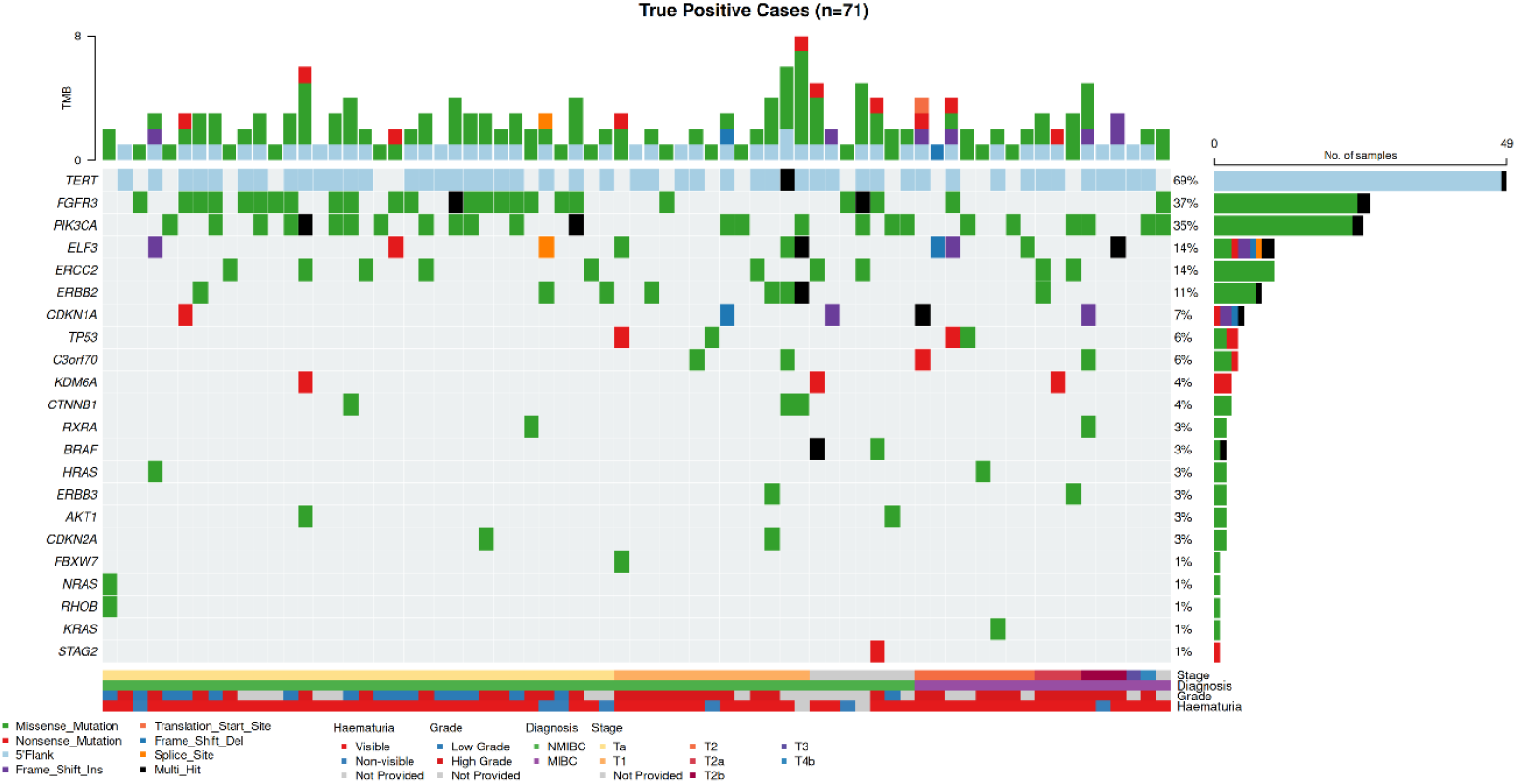
Mutational landscape of true positive bladder cancer cases detected by GALEAS™ Bladder. Oncoplot showing diagnostic mutations in 71 bladder cancer cases. Each column represents a patient sample; each row represents a gene. Top panel shows variant allele frequency (VAF) distribution. Green squares indicate missense mutations, red squares indicate frameshift mutations, black squares indicate nonsense mutations, and orange/blue squares indicate other mutation types. *TERT* promoter mutations were most frequent (69%), followed by *FGFR3* (38%), *PIK3CA* (35%), *ELF3* (14%), and *ERCC2* (14%). Right panel shows mutation frequency across the cohort. Bottom annotation bars indicate haematuria type (visible/non-visible/not provided), clinical diagnosis (NMIBC/MIBC), tumour stage (Ta/T1/T2+), and tumour grade (high grade/low grade). Multiple mutations per sample are common, reflecting the multi-gene panel approach.

