## Supplemental Methods and Figures for "GALEAS™ Bladder Demonstrates High Sensitivity and Specificity for Detecting Bladder Cancer: Real-World Multicentre Data from UK NHS Haematuria Clinics"

**Disclosures:** DGW and RTB co-developed the GALEAS™ Bladder urine test with Nonacus Ltd (UK). RTB is a paid consultant for Cystotech ApS (Denmark) and an unpaid charity trustee for Action Bladder Cancer UK (UK). LS, RKH, SC, RS, MP, MS and VH are employees of Nonacus.

#### Supplemental Methods

FASTQ files are processed using our proprietary bioinformatics algorithm that leverages error suppression technology, with downstream custom algorithms for automated variant classification and reporting specific to BC detection. Annotated variants are evaluated by the GALEAS™ Bladder classifier, which identifies diagnostic mutations in bladder cancer-associated genes including *TERT* promoter hotspots and tier I/II oncogenic variants in genes such as *FGFR3*, *PIK3CA*, *ELF3*, and *ERCC2*. Samples were classified as positive for bladder cancer if at least one mutation was detected.

#### Statistical methods

##### Study Design and Population

This prospective diagnostic accuracy study enrolled consecutive patients presenting with haematuria at 7 NHS sites in the UK between October 2024 and June 2025. Eligibility criteria included adults aged ≥18 years referred for investigation following the detection of either visible haematuria (VH) or non-visible haematuria (NVH), with the ability to provide informed consent and a voided urine sample prior to cystoscopy. Exclusion criteria comprised a prior diagnosis of urothelial cancer, inability to provide an adequate urine sample, or unwillingness or inability to undergo standard haematuria investigations

The study was conducted and reported in accordance with the STARD 2015 guidelines for diagnostic accuracy studies.

Based on prior performance data for GALEAS™ Bladder [8] (87.3% (95% confidence interval [CI] 81.2-92.0%) at specificity of 84.8% (95% CI 79.9-89.0%)), this sample size was calculated to provide a precision of ±5% for sensitivity and ±2% for specificity, using 95% exact binomial confidence intervals.

##### Diagnostic Performance Metrics

Sensitivity, specificity, positive predictive value (PPV), and negative predictive value (NPV) were calculated from 2×2 contingency tables. Wilson score 95% confidence intervals were calculated for all proportions, as recommended for binomial data with small sample sizes or proportions approaching 0% or 100%.

##### Likelihood Ratios

The positive likelihood ratio (LR+) was calculated as sensitivity/(1−specificity), and the negative likelihood ratio (LR-) as (1-sensitivity)/specificity. Confidence intervals for likelihood ratios were calculated using the log transformation method:

$$SE(\ln LR+) = \sqrt{1/TP - 1/(TP+FN) + 1/DP - 1/(DP+TN)}$$

$SE(\ln LR-) = \sqrt{(1/FN - 1/(TP+FN) + 1/TN - 1/(DP+TN))}$

TP=Classified a s likely bladder cancer by detection of a bladder cancer mutation and pathology confirmed bladder cancer

DP=Classified as likely bladder cancer by detection of a bladder cancer mutation but reported as No Cancer by cystoscopy.

FN= Classified as no bladder cancer due to not detecting bladder cancer mutations but pathology confirmed bladder cancer

TN= Classified as no bladder cancer due to not detecting bladder cancer mutations and reported as No Cancer Clinically

**Diagnostic Odds Ratio**

The diagnostic odds ratio (DOR) was calculated as (TP×TN)/(DP×FN), representing the odds of a positive test result in patients with cancer relative to those without. Confidence intervals were calculated using the log transformation method:

$SE(\ln DOR) = \sqrt{(1/TP + 1/TN + 1/DP + 1/FN)}$

**Number Needed to Test**

The number needed to test (NNT) was calculated as the reciprocal of PPV (1/PPV), representing the number of GALEAS-positive patients requiring cystoscopy to detect one cancer case. NNT confidence intervals were derived from the inverse of PPV confidence limits. Cystoscopy reduction was calculated as the proportion of test-negative patients who could potentially defer immediate cystoscopy.

**Subgroup Analyses**

Diagnostic performance was evaluated within pre-specified clinically relevant subgroups: tumour stage (Ta, T1, T2+), tumour grade (high-grade, low-grade), and haematuria type (visible, non-visible). Subgroup analyses were exploratory; no adjustment for multiple comparisons was performed, and results should be interpreted accordingly. The probability of at least one false-positive finding across multiple subgroups exceeds the nominal 5% significance level.

**Variant Allele Frequency Analysis**

VAF distributions were compared between true positive (cancer-detected) and discordant positive (no cancer via cystoscopy) samples using violin plots with overlaid data points. Summary statistics (median, interquartile range) were calculated for each classification group and by diagnosis category.

#### Statistical Software

All analyses were performed in R (version 4.5.0) using the tidyverse, gt, and gtsummary packages. Oncoplots were generated with maftools. Wilson score confidence intervals were implemented using the standard formula. Figures were generated using ggplot2 with colorblind-accessible palettes (Okabe-Ito).

#### Supplemental Figures

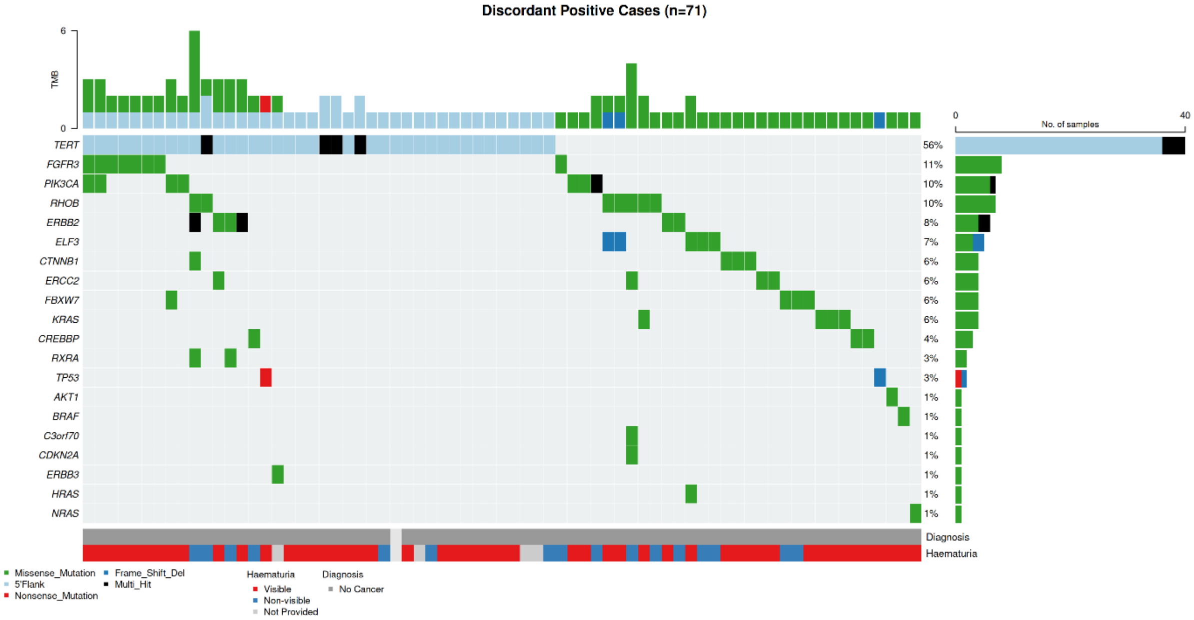


**Supplementary Figure S1. Mutational profile of discordant positive cases (n=71)**.

Oncoplot of mutations detected by GALEAS™ Bladder in cystoscopy-negative patients. Each column represents a patient; rows represent genes. Top panel: VAF distribution (0.2-72%). Colours indicate mutation types (green=missense, red=frameshift, black=nonsense, blue=other). *TERT* was most frequently mutated (56%), followed by *FGFR3* (11%), *PIK3CA* (10%), and *RHOB* (10%). Right panel shows mutation frequencies. Bottom annotations show haematuria type and diagnosis.

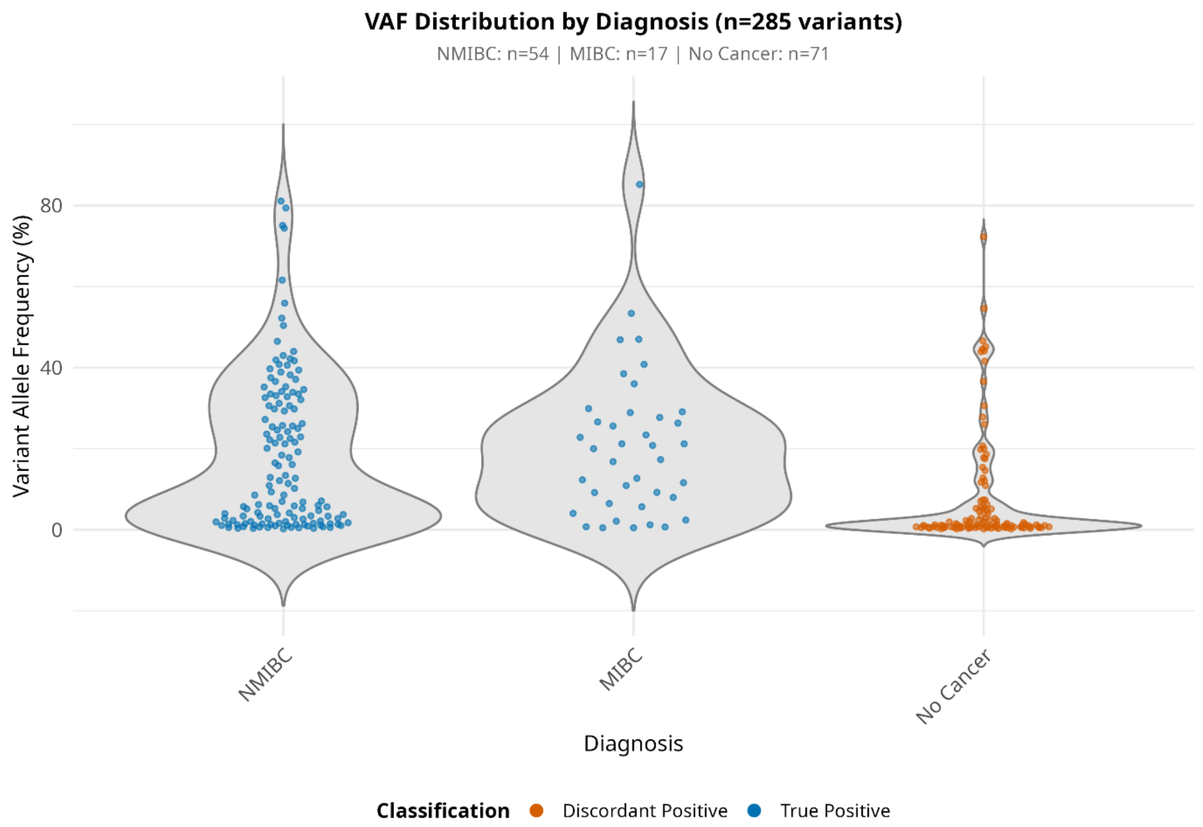


**Supplementary Figure S2A. Variant allele frequency distribution by clinical diagnosis.** Violin plots showing VAF distributions for all diagnostic mutations detected by GALEAS Bladder, stratified by clinical diagnosis. Each point represents a single somatic variant. MIBC (blue, n=17 patients) and NMIBC (blue, n=54 patients) both show broad VAF distributions (0.2-85%), with variants spanning low to high frequencies. No Cancer (orange, n=71 discordant positive patients) demonstrates a bimodal distribution: 40% of variants at VAF <1% and 60% at VAF >=1% 1-72.3% (range 0.2-72.3%). Violin width represents variant density at each VAF level.

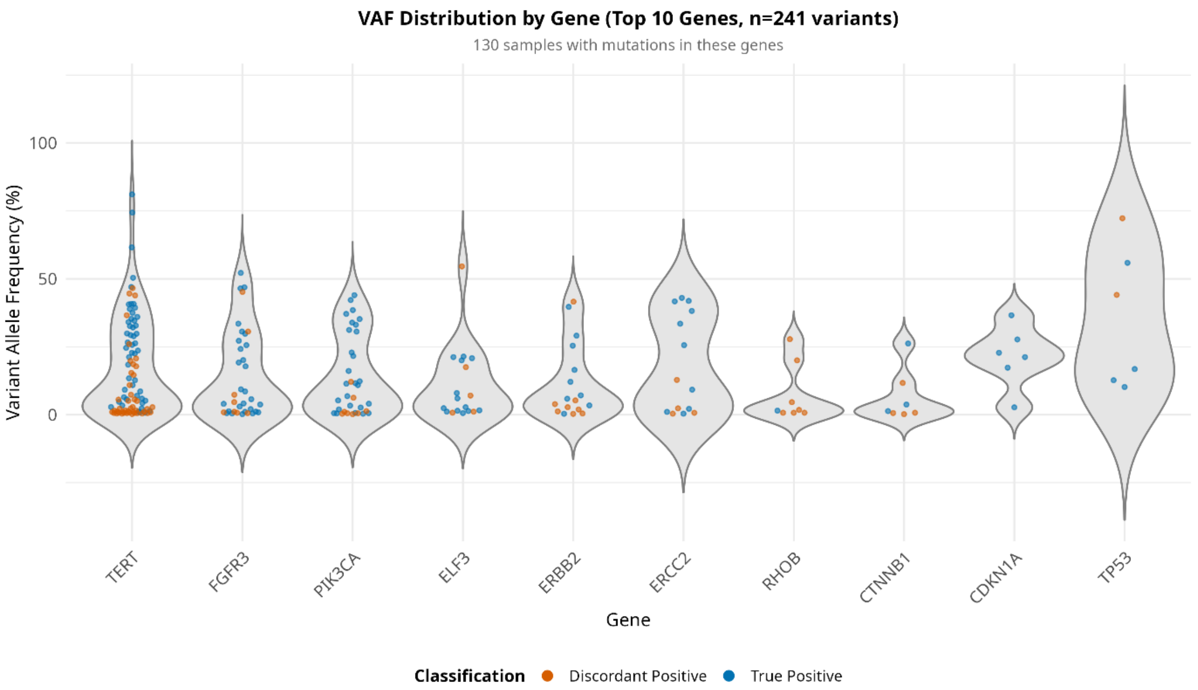


**Supplementary Figure S2B. Variant allele frequency distribution by gene and classification.****

Violin plots showing VAF distributions for the ten most frequently mutated genes, with variants color-coded by classification: TP (true positive, purple) for variants in cystoscopy-confirmed bladder cancer patients; DP (discordant positive, teal) for variants in patients without confirmed cancer. *TERT*, *FGFR3*, *PIK3CA*, *ELF3*, *ERBB2*, and *ERCC2* show overlapping VAF distributions between TP and FP cases, with both groups containing high-VAF mutations, suggesting some FP cases may represent occult malignancies. The presence of high-VAF driver mutations in DP cases supports the hypothesis that molecular testing detects early disease below cystoscopic detection thresholds.

#### Supplemental Tables

##### Table S1

*Counts Per Site*

| Site | N | Bladder Cancer | No Bladder Cancer |
|---|---|---|---|
| Site 1 | 455 | 36 | 419 |
| Site 2 | 121 | 12 | 109 |
| Site 3 | 85 | 4 | 81 |
| Site 4 | 84 | 7 | 77 |
| Site 5 | 78 | 4 | 74 |
| Site 6 | 74 | 6 | 68 |
| Site 7 | 67 | 8 | 59 |
| Overall | 964 | 77 | 887 |

##### Table S2A

*S2A - VAF distribution by GALEAS Bladder Classification*

| Classification | n_variants | n_samples | min_vaf | q25_vaf | median_vaf | q75_vaf | max_vaf | mean_vaf |
|---|---|---|---|---|---|---|---|---|
| DP (Discordant Positive) | 110 | 71 | 0.20 | 0.64 | 1.21 | 6.09 | 72.30 | 7.72 |
| TP (True Positive) | 175 | 71 | 0.22 | 2.67 | 12.70 | 30.25 | 85.20 | 18.71 |

##### Table S2B

*S2B - VAF distribution by Clinical Diagnosis*

| Diagnosis | n_variants | n_samples | median_vaf | q25_vaf | q75_vaf |
|---|---|---|---|---|---|
| NMIBC | 136 | 54 | 11.15 | 2.12 | 31.42 |
| MIBC | 39 | 17 | 20 | 7.21 | 28.30 |
| No Cancer | 110 | 71 | 1.21 | 0.64 | 6.09 |

### Table 3A

S3A Summary of Sensitivity, Specificity, PPV and NPV

| **Group** | **n_total** | **n_cancer** | **n_positive** | **tp** | **dp** | **tn** | **fn** | **sensitivity** | **sensitivity_lower** | **sensitivity_upper** | **specificity** | **specificity_lower** | **specificity_upper** | **ppv** | **ppv_lower** | **ppv_upper** | **npv** | **npv_lower** | **npv_upper** |
|---|---|---|---|---|---|---|---|---|---|---|---|---|---|---|---|---|---|---|---|
| Overall | 964 | 77 | 142 | 71 | 71 | 816 | 6 | 92.20 | 84 | 96.40 | 92 | 90 | 93.60 | 50 | 41.90 | 58.10 | 99.30 | 98.40 | 99.70 |
| Visible Haematuria | 575 | 68 | 113 | 62 | 51 | 456 | 6 | 91.20 | 82.10 | 95.90 | 89.90 | 87 | 92.30 | 54.90 | 45.70 | 63.70 | 98.70 | 97.20 | 99.40 |
| Non-visible Haematuria | 294 | 7 | 23 | 7 | 16 | 271 | 0 | 100 | 64.60 | 100 | 94.40 | 91.10 | 96.50 | 30.40 | 15.60 | 50.90 | 100 | 98.60 | 100 |

### Table 3B

S3B Summary of Likelihood Ratios, Diagnostic Ratio, Referral rate, Number Needed to Treat and Cystoscopy Reduction

| Group | n_total | n_cancer | n_positive | tp | dp | tn | fn | lr_pos | lr_pos_lower | lr_pos_upper | lr_neg | lr_neg_lower | lr_neg_upper | dor | dor_lower | dor_upper | referral_rate | nnt | nnt_lower | nnt_upper | cysto_reduction |
|---|---|---|---|---|---|---|---|---|---|---|---|---|---|---|---|---|---|---|---|---|---|
| Overall | 964 | 77 | 142 | 71 | 71 | 816 | 6 | 11.52 | 9.13 | 14.53 | 0.09 | 0.04 | 0.18 | 136 | 57.10 | 323.90 | 14.70 | 2 | 1.72 | 2.39 | 85.30 |
| Visible Haematuria | 575 | 68 | 113 | 62 | 51 | 456 | 6 | 9.06 | 6.92 | 11.88 | 0.10 | 0.05 | 0.21 | 92.40 | 38.10 | 224.20 | 19.70 | 1.82 | 1.57 | 2.19 | 80.30 |
| Non-visible Haematuria | 294 | 7 | 23 | 7 | 16 | 271 | 0 | 17.94 | 11.14 | 28.88 | 0 | | | Inf | | | 7.80 | 3.29 | 1.97 | 6.41 | 92.20 |

Table S3C

*Metrics Description*

| Field | Description |
|---|---|
| Group | Stratification group (Overall, Visible Haematuria, Non-visible Haematuria) |
| n_total | Total analyzable samples in this group |
| n_cancer | Number of confirmed bladder cancer cases (TP + FN) |
| n_positive | Number of GALEAS™ positive results (TP + DP) |
| tp | True Positives - cancer cases correctly identified with a positive GALEAS Bladder Mutation |
| dp | Discordant Positives - non-cancer cases with positive GALEAS Bladder Mutation |
| tn | True Negatives - non-cancer cases correctly identified as negative |
| fn | False Negatives - cancer cases missed, No GALEAS Bladder Mutation detected |
| sensitivity | Proportion of cancer cases detected (TP / n_cancer × 100) |
| sensitivity_lower | Lower bound of 95% Wilson CI for sensitivity |
| sensitivity_upper | Upper bound of 95% Wilson CI for sensitivity |
| specificity | Proportion of non-cancer cases correctly identified (TN / (TN+DP) × 100) |
| specificity_lower | Lower bound of 95% Wilson CI for specificity |
| specificity_upper | Upper bound of 95% Wilson CI for specificity |
| ppv | Positive Predictive Value - probability positive test = cancer (TP / n_positive × 100) |
| ppv_lower | Lower bound of 95% Wilson CI for PPV |
| ppv_upper | Upper bound of 95% Wilson CI for PPV |
| npv | Negative Predictive Value - probability negative test = no cancer (TN / (TN+FN) × 100) |

| | |
|---|---|
| npv_lower | Lower bound of 95% Wilson CI for NPV |
| npv_upper | Upper bound of 95% Wilson CI for NPV |
| lr_pos | Positive Likelihood Ratio = Sensitivity / (1 - Specificity). |
| lr_pos_lower | Lower bound of 95% CI for LR+ (log method) |
| lr_pos_upper | Upper bound of 95% CI for LR+ (log method) |
| lr_neg | Negative Likelihood Ratio = (1 - Sensitivity) / Specificity. |
| lr_neg_lower | Lower bound of 95% CI for LR- (log method) |
| lr_neg_upper | Upper bound of 95% CI for LR- (log method) |
| dor | Diagnostic Odds Ratio = (TP × TN) / (DP × FN). |
| dor_lower | Lower bound of 95% CI for DOR (log method) |
| dor_upper | Upper bound of 95% CI for DOR (log method) |
| referral_rate | Percentage of patients requiring cystoscopy referral (n_positive / n_total × 100) |
| nnt | Number Needed to Treat - cystoscopies per cancer detected (n_positive / TP = 1/PPV) |
| nnt_lower | Lower bound of 95% CI for NNT (1 / PPV_upper) |
| nnt_upper | Upper bound of 95% CI for NNT (1 / PPV_lower) |
| cysto_reduction | Percentage of patients who could avoid cystoscopy (100 - referral_rate) |

#### Table S4A

*S4A - Mutation Summary Counts*

| Metric | Value |
|---|---|
| Total positive samples | 142 |
| Total variants | 285 |
| Mean variants/sample | 2 |
| Min VAF | 0.0020 |
| Max VAF | 0.85 |
| Median VAF | 0.06 |
| Samples with >1 mutation | 76/142 (54%) |

#### Table S4B

*S4B - GALEAS Bladder Positive calls across diagnosis, stage and grade*

| Gene | Overall (N=142) | NMIBC (N=54) | MIBC (N=17) | No Cancer (N=71) | Ta (N=34) | T1 (N=13) | T2+ (N=16) | High Grade (N=35) | Low Grade (N=16) |
|---|---|---|---|---|---|---|---|---|---|
| TERT | 89 | 38 | 11 | 40 | 24 | 9 | 11 | 26 | 9 |
| FGFR3 | 34 | 24 | 2 | 8 | 20 | 1 | 1 | 7 | 11 |
| PIK3CA | 32 | 19 | 6 | 7 | 13 | 3 | 5 | 8 | 9 |
| ELF3 | 15 | 6 | 4 | 5 | 3 | 3 | 4 | 5 | 1 |
| ERBB2 | 14 | 7 | 1 | 6 | 3 | 4 | 1 | 5 | 0 |
| ERCC2 | 14 | 8 | 2 | 4 | 5 | 1 | 2 | 7 | 0 |
| RHOB | 8 | 1 | 0 | 7 | 1 | 0 | 0 | 0 | 1 |
| CTNNB1 | 7 | 3 | 0 | 4 | 1 | 2 | 0 | 0 | 1 |
| TP53 | 6 | 2 | 2 | 2 | 0 | 2 | 2 | 2 | 0 |
| C3orf70 | 5 | 2 | 2 | 1 | 0 | 2 | 2 | 3 | 0 |
| CDKN1A | 5 | 3 | 2 | 0 | 1 | 1 | 2 | 3 | 1 |
| FBXW7 | 5 | 1 | 0 | 4 | 0 | 1 | 0 | 1 | 0 |
| KRAS | 5 | 0 | 1 | 4 | 0 | 0 | 1 | 1 | 0 |
| RXRA | 4 | 1 | 1 | 2 | 1 | 0 | 1 | 2 | 0 |
| AKT1 | 3 | 2 | 0 | 1 | 1 | 0 | 0 | 1 | 1 |
| BRAF | 3 | 2 | 0 | 1 | 0 | 0 | 0 | 1 | 0 |
| CDKN2A | 3 | 2 | 0 | 1 | 1 | 1 | 0 | 2 | 0 |
| CREBBP | 3 | 0 | 0 | 3 | 0 | 0 | 0 | 0 | 0 |
| ERBB3 | 3 | 1 | 1 | 1 | 0 | 1 | 1 | 2 | 0 |
| HRAS | 3 | 1 | 1 | 1 | 1 | 0 | 1 | 2 | 0 |
| KDM6A | 3 | 2 | 1 | 0 | 1 | 0 | 1 | 2 | 0 |
| NRAS | 2 | 1 | 0 | 1 | 1 | 0 | 0 | 0 | 1 |
| STAG2 | 1 | 1 | 0 | 0 | 0 | 0 | 0 | 1 | 0 |
| Any (n) | 142 | 54 | 17 | 71 | 34 | 13 | 16 | 35 | 16 |
| Sensitivity (%) | 92.20 | 90 | 100 | — | 85 | 100 | 100 | 97.20 | 84.20 |
| Specificity (%) | 92 | 92 | 92 | 92 | 92 | 92 | 92 | 92 | 92 |
